# An Automated, Pathologist-free Gleason Grade Stratifies Disease-free Interval Comparably to Expert Grading from a Single Out-of-distribution Slide

**DOI:** 10.64898/2026.06.22.26356247

**Authors:** Joshua L. Ebbert, Jared Szymansky, Anthony Perry, Dennis Della Corte

## Abstract

Automated Gleason grading now matches expert pathologists on the cohorts where systems are developed and tuned, but deployment-relevant gaps remain: whether an automated grade, applied without site-specific tuning or pathologist oversight, stratifies outcome comparably to expert grading on slides from unseen institutions and in cross-specimen applications. We tested this for disease-free interval (DFI), a curated recurrence endpoint. A production gland-level prostate diagnostic (PathTools Prostate v11.0) was applied frozen and uncalibrated to 298 diagnostic whole-slide images from 274 TCGA-PRAD radical-prostatectomy patients, a cohort outside its development distribution and needle-core-biopsy training data, contributed by 25 source sites under heterogeneous digitization; tissue was detected automatically with no expert region annotation. From the output we derived an ISUP grade group and continuous high-grade content, and evaluated each grade as a standalone predictor of DFI (24 events) by Harrell’s c-index with 95% bootstrap confidence intervals, a paired between-method bootstrap, and Kaplan-Meier curves with the log-rank test. The automated grade reproduced the clinical grade group at quadratic-weighted κ = 0.62 (95% CI 0.53–0.70; 48% exact, 86% within one group), within the expert inter-observer range. As the sole predictor it stratified recurrence (log-rank *p* = 0.022; c-index 0.69, 95% CI 0.58–0.79), and the continuous high-grade fraction was robustly prognostic (hazard ratio 1.37 per SD, *p* = 0.029; c-index 0.71, 0.61–0.81). Standalone discrimination was not statistically separable from the clinical grade (c-index 0.78, 0.69–0.86; paired Δ c-index spanning zero), and in a joint model the automated grade added nothing beyond it, consistent with both measuring a shared morphological axis. From a single out-of-distribution slide with no pathologist oversight, the automated grade provides standalone recurrence stratification not statistically separable from whole-gland expert grading, demonstrating robust generalizability beyond training data; reported as a continuous high-grade fraction, it offers reproducible, expert-free, grade-equivalent risk stratification for harmonizing large archival or genomically-profiled cohorts.

## 1. INTRODUCTION

The Gleason system, recast by the 2014 ISUP consensus into five ordinal grade groups, remains the single most powerful routinely available prognostic variable in localized prostate cancer [1, 2]. Its weakness is reproducibility. Grading is a subjective morphological judgement, and inter-observer agreement, even among specialist urological pathologists, sits only at substantial levels (quadratic-weighted κ ≈ 0.56 to 0.70), falling to barely moderate (κ ≈ 0.44) among general pathologists, with the intermediate grades and cribriform morphology the chief sources of disagreement [3, 4]. Because grade drives treatment selection, this variability propagates directly into care with concrete clinical consequences [5-7].

Deep learning is the proposed remedy. Systems trained to grade prostate biopsies and prostatectomies reach or exceed general-pathologist accuracy and approach expert panels [8-10], and the PANDA challenge demonstrated pathologist-level grading across cross-continental validation cohorts [11]. Two gaps separate these results from deployment. The first is generalization. Histology models routinely learn site- and scanner-specific signatures rather than biology, so accuracy measured on internally drawn or co-acquired validation data overstates real-world performance: deep networks can identify the contributing TCGA site from the image alone, and this leakage inflates apparent accuracy [12]. An honest test uses data from institutions and scanners entirely outside the development distribution, scored without site-specific recalibration. A system that needs a curated, site-specific calibration set before deployment cannot straightforwardly extend access to institutions that lack the annotated cases, the pathologist time, or the scanner infrastructure to supply them, although this requirement is common even in leading commercial systems [13]. Generalization also includes cross-specimen performance, which represents a significant domain shift problem [14-16] and can be limited by factors such as grading conventions. The threshold and criteria for assigning secondary Gleason patterns differs, for example, between needle core biopsies and resections. The second gap is the pathologist-in-the-loop assumption. Most validations report agreement with an expert reference, presupposing that an expert is available to grade, or to annotate the region the model grades. The scenario that would actually extend access, fully automated grade assignment from a raw slide, is rarely isolated and almost never evaluated for prognostic value rather than concordance alone.

We address both gaps directly. A production gland-level diagnostic, PathTools Prostate v11.0, trained on needle core biopsy data to detect tumors, grade glands, and segment cribriform and perineural morphology end-to-end from a whole slide with no human region annotation, is applied frozen and uncalibrated to the TCGA-PRAD radical-prostatectomy cohort [17]. TCGA-PRAD is a stringent out-of-distribution test: its slides come from 25 tissue source sites digitized under heterogeneous conditions, none in the model’s development, and the cohort carries a curated disease-free interval endpoint [18]. The question is not whether the AI agrees with pathologists, but whether the automated grade, assigned with no pathologist and no tuning and used as the sole available predictor, stratifies disease-free interval and new tumor events as well as the expert grade. This standalone comparison, not the incremental value of one grade over the other, is the deployment-relevant question, because in the settings that motivate the work only one grade is available. It determines whether such tools can substitute for expert grading where none exists, for example to harmonize grade across large archival and genomically-profiled prostatectomy cohorts (such as those characterized by genomic classifiers like Decipher [19, 20]) for which uniform central re-grading was never performed.

## 2. MATERIALS AND METHODS

### 2.1 Cohort and outcome

We assembled all open-access TCGA-PRAD diagnostic (-DX, FFPE) whole-slide images with a recorded disease-free outcome: 298 slides from 274 patients, downloaded from the NCI Genomic Data Commons (Grossman et al. 2016). The endpoint was disease-free interval (DFI), a curated recurrence and progression endpoint from the TCGA Pan-Cancer Clinical Data Resource [18], obtained via cBioPortal (prad_tcga_pan_can_atlas_2018) [21]: 24 events among 274 patients (∼9 %), median follow-up ∼32 months. Pathologic T/N stage and age came from the same source. PSA and surgical-margin status are not available in TCGA. Most patients (267 of 274) are represented by a single diagnostic slide, whereas the clinical grade was assigned from the full prostatectomy specimen (Section 4).

### 2.2 Automated diagnostic: No fine-tuning or pathologist oversight

Every slide was processed by PathTools Prostate v11.0 exactly as deployed clinically, with no retraining, fine-tuning, color normalization, or recalibration to TCGA. Tissue was detected automatically (Otsu thresholding on image saturation); no expert region boxes or annotations were used at any stage. The pipeline ingests whole-slide images and outputs tumor detection, Gleason grade prediction, and segmentation of cribriform pattern and perineural invasion. We treat the system as a fixed black box; further architectural and model details are not disclosed. From each slide’s diagnostic output, we derived an ISUP grade group from the area-ranked primary and secondary patterns, as a pathologist assigns it from dominant morphology.

### 2.3 Predictors

From the AI diagnostic we derived, per patient (area-pooled aggregation): the automated grade group; the continuous high-grade fraction (% Gleason 4+5 of tumor), which retains the 3-versus-4 gradient that grade-group discretization discards; and a soft per-tile grader-softmax composition (tumor-normalized mean grade and % Gleason 4+5), available for a representative subset (n = 205, 22 events). The comparator was the clinical Gleason grade group from the original TCGA report. Pathologic T/N stage, age, and AI cribriform extent were used as additional covariates.

### 2.4 Statistical analysis

Grade-group concordance between the automated and clinical grades was summarized by a quadratic-weighted Cohen’s κ (bootstrap 95 % CI), exact and within-one-group agreement, and the per-clinical-group mean automated grade. The primary prognostic analysis evaluated each grade as a standalone predictor of new tumor events by Harrell’s c-index, reported with a 95 % confidence interval from a 2,000-replicate bootstrap; the paired between-method difference in c-index was estimated by scoring both grades on the same resample at each replicate, so its interval reflects their correlation. Kaplan-Meier estimation with the log-rank test (grade-group strata and median splits) provided the visualization, and univariate Cox proportional-hazards models the hazard ratios. A secondary joint Cox model containing both grades, with bidirectional likelihood-ratio tests, characterized their redundancy (whether either grade adds information given the other). A multivariable Cox model tested whether pathologic stage adds to the automated grade. Given 24 events, a ∼10-events-per-covariate ceiling was enforced; the study is powered to exclude a large standalone gap, not a moderate one, and results are reported with that limit foregrounded. Analyses used lifelines, scikit-learn, scipy, and numpy; code and the de-identified feature table are released (Section 6).

### 2.5 Sensitivity analysis

TCGA-PRAD supports three distinct recurrence endpoints: the disease-free interval (DFI) used here, the progression-free interval (PFI; *does not require disease-free baseline*), and true biochemical (PSA) recurrence from the legacy clinical release. We repeated the entire analysis on all three (Supplementary Information).

## 3. RESULTS

Figure 1 shows representative resections with the automated overlay across the cohort’s prognostic range: high-grade disease demonstrating Gleason pattern 5 (shown) and cribriform architecture (not shown) across three diagnostic slides that recurred (A); the systematic low-end over-call on a clinically indolent Gleason 3+3 tumor (B); an indolent low-grade tumor disease-free at 14 years (C); and diffuse high-grade tumor with rapid recurrence (D).

**Figure 1.**
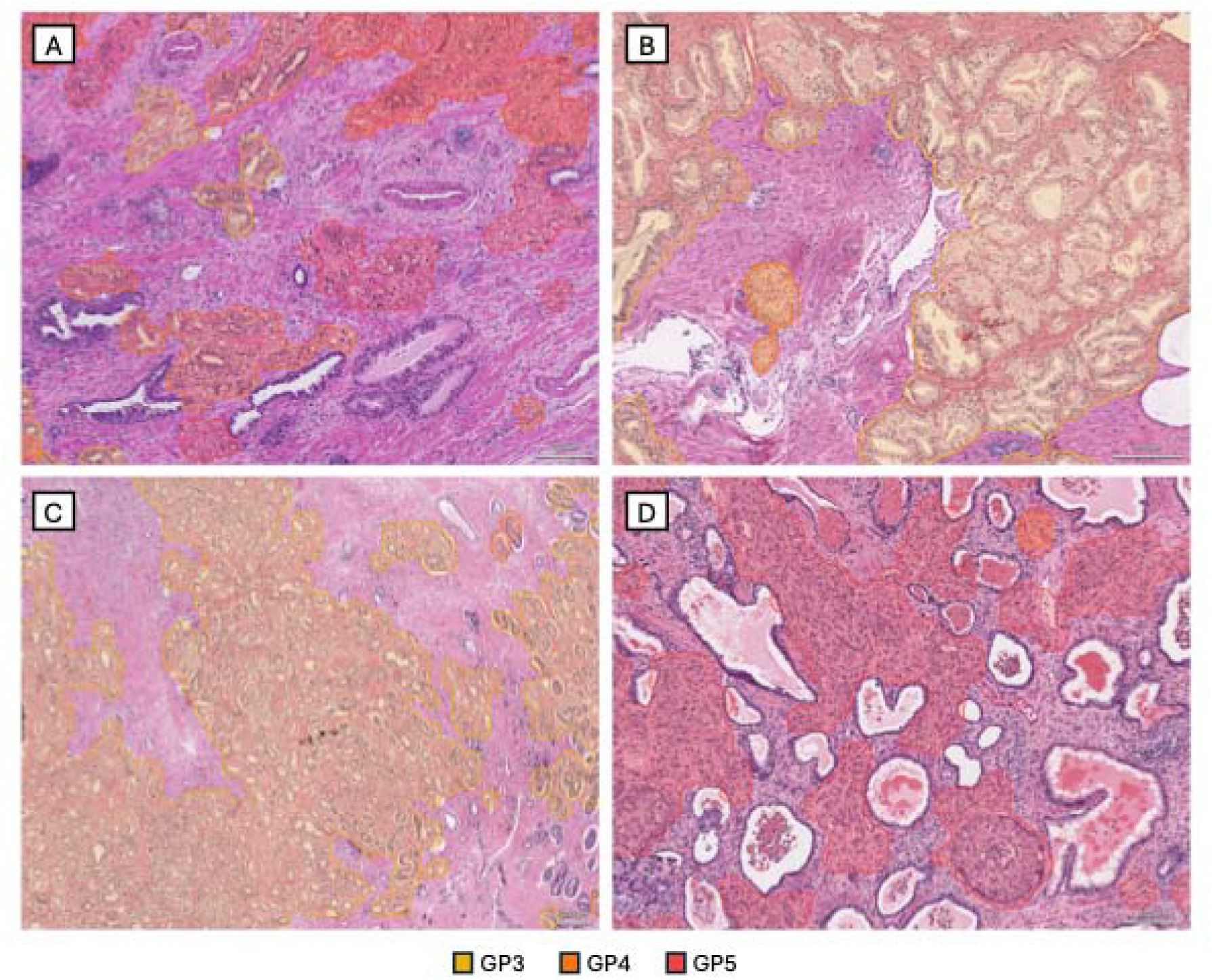
Representative TCGA-PRAD resections with the automated PathTools Prostate v11.0 Gleason overlay (no expert region annotation). (A) Concordant high grade (clinical and AI grade group 5); recurred at 25 months (pT3b). (B) Pathologist Gleason 3+3 (grade group 1) upgraded to grade group 2 by the AI on trace pattern-4 calls, the systematic low-end over-call under acquisition domain shift; clinically indolent (disease-free at 86 months). (C) Indolent low-grade tumor, disease-free at 165 months. (D) Diffuse high-grade (Gleason 4+5) tumor, recurrence at 4 months (pT4). Gleason pattern 3 yellow, pattern 4 orange, pattern 5 red; scale bar 200 µm.

### 3.1 Grade group concordance

Across the 272 patients with both grades, the automated grade group reproduced the clinical grade group at quadratic-weighted κ = 0.62 (95 % CI 0.53 to 0.70), with 48 % exact and 86 % within-one-group agreement (Figure 2A), within the inter-observer range reported for specialist urological pathologists [3] and broadly consistent with published expert-validated deep-learning graders [8, 9]. The mean automated grade rose monotonically across clinical groups GG1 to GG5 (2.10, 2.60, 3.01, 3.88, 4.33; Figure 2B). The discrepancies are systematic and concentrate at the extremes. At the low end the AI over-calls: clinical GG1 (Gleason 3+3) is usually read as GG2 (23 of 30). At the high end, the AI under-calls a subset of high-grade tumors: most clinical GG5 are still read as GG5 (28 of 42, with a healthy median 22 % genuine pattern-5 area), but 14 of 42 drop to GG3. The model detects genuine pattern 5 confidently when it is morphologically present; the drops occur in cribriform-rich tumors with little solid pattern 5, where the high grade is now carried by pattern 4.

**Figure 2.**
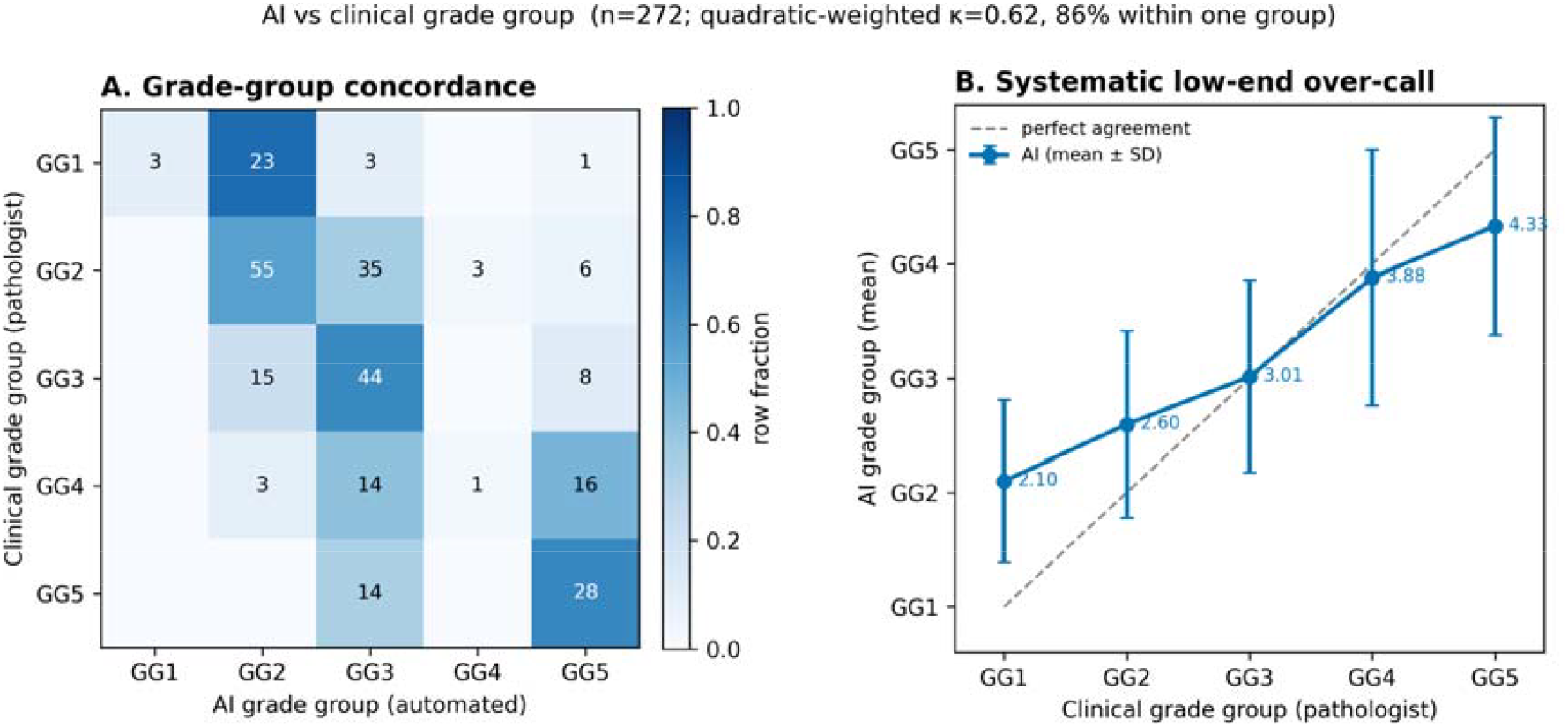
The automated grade group reproduces the pathologist grade group on TCGA-PRAD (n = 272). (A) Confusion matrix of clinical (pathologist) versus AI grade group, row-fraction shaded, cell counts shown. (B) Mean AI grade group by clinical grade group (±SD); the AI grade rises monotonically but over-calls the low end (clinical GG1 read as GG2). Quadratic-weighted κ = 0.62; 86 % within one group.

### 3.2 Recurrence stratification

As the sole predictor, the automated grade group stratified disease-free survival by the log-rank test (*p* = 0.022; Figure 3B), though its univariate Cox hazard ratio was borderline at this event count (1.23 per group, *p* = 0.078); the continuous AI %Gleason 4+5 was robustly prognostic (hazard ratio 1.37 per SD, *p* = 0.029; median-split log-rank *p* = 0.001; Figure 3C). The Kaplan-Meier separation by the automated grade is visually super-imposable on the separation by the clinical grade (log-rank *p* < 0.001; Figure 3A).

**Figure 3.**
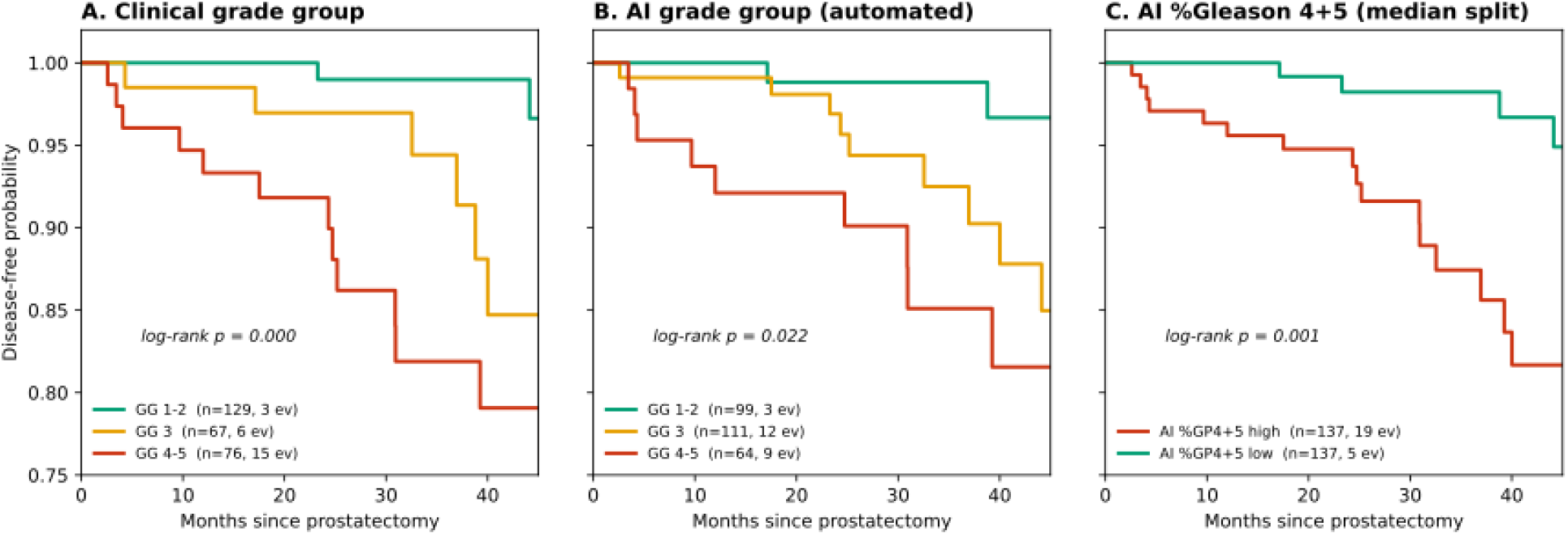
The automated grade carries the same disease-free-survival signal as the expert grade. Kaplan-Meier disease-free survival stratified by (A) clinical grade group, (B) the automated AI grade group (same strata; visually super-imposable on A), and (C) AI %Gleason 4+5 at the median split. Log-rank p shown per panel; ev = recurrence events.

Quantified by standalone discrimination with matched confidence intervals (Figure 4A), the clinical Gleason grade reached c-index 0.78 (95 % CI 0.69 to 0.86), the automated grade group 0.69 (0.58 to 0.79), and the continuous AI %Gleason 4+5 0.71 (0.61 to 0.81). The paired between-method difference covered zero in every case (AI grade group Δ c-index 0.09, 95 % CI −0.01 to +0.20; AI %Gleason 4+5 Δ c-index 0.07, 95 % CI −0.02 to +0.16), so the automated and expert grades were not statistically separable as standalone recurrence predictors at this event count, although the point estimates favored the expert grade. The continuous high-grade fraction was the closest proxy and exceeded the discretized AI grade group.

**Figure 4.**
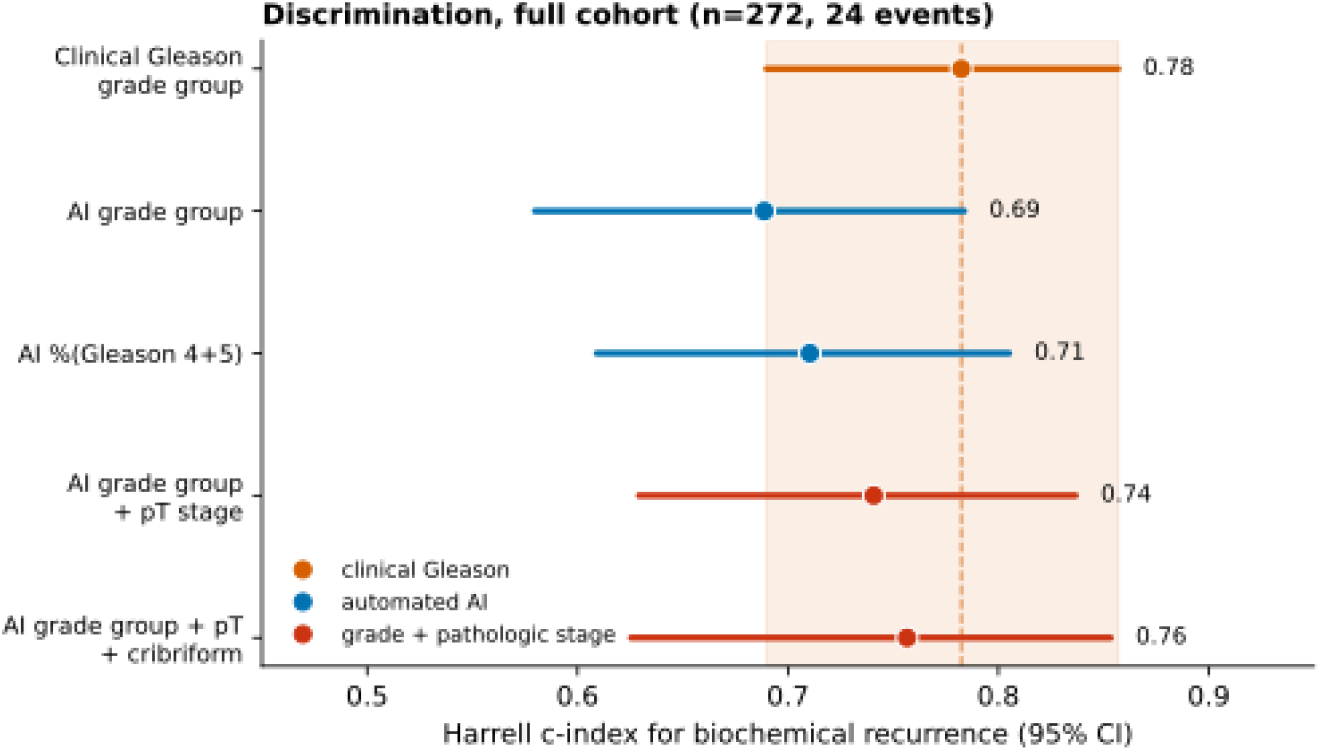
Standalone prognostic discrimination of clinical versus automated predictors, with confidence intervals. Harrell c-index for new tumor events (point estimate and 95 % bootstrap CI); the shaded band marks the clinical grade’s CI so overlap is direct. Full cohort (n = 272, 24 events): the automated grade group and continuous AI %Gleason 4+5 overlap the clinical grade; the automated grade plus pathologic T stage (± cribriform) reaches c-index 0.74. Every AI-versus-clinical paired difference has a 95 % CI covering zero.

### 3.3 Prognostic independence

A joint Cox model containing both grades quantifies their relationship. The automated grade added no information beyond the clinical grade (likelihood-ratio *p* = 0.50; joint c-index 0.78, unchanged from the clinical grade alone). We could not detect an independent contribution of the AI grade beyond the clinical grade, consistent with the two measuring a shared morphological axis, though our power to detect moderate independent contributions is limited at this event count. The clinical grade retained a small increment over the AI grade (likelihood-ratio *p* = 0.005). The per-tile soft composition behaved identically to the hard features: on the soft subset (n = 205, 22 events) AI soft mean grade and AI soft %Gleason 4+5 reached c-index 0.69 and 0.70, each redundant with the clinical grade (likelihood-ratio *p* > 0.2). Among AI morphometry, only cribriform extent carried independent univariate signal (hazard ratio 1.21 per SD, *p* = 0.042); tumor area, focality, gland size, age and nodal status added nothing. The one covariate that added beyond grade was pathologic T stage (univariate hazard ratio 1.90, *p* = 0.014; in a joint model with the AI grade, hazard ratio 2.19, *p* = 0.016): the automated grade plus pT stage reached c-index 0.74 (95 % CI 0.64 to 0.83; Figure 4A), approaching post-prostatectomy clinical nomograms such as CAPRA-S [22] even though TCGA lacks the PSA and surgical-margin inputs those nomograms use.

#### Sensitivity analysis

TCGA-PRAD supports three distinct recurrence endpoints. The sensitivity analysis findings are endpoint-robust: grade-group concordance is unchanged (quadratic-weighted κ = 0.62), the automated grade stratifies recurrence on every endpoint (log-rank p = 0.02–0.03), every automated-versus-clinical paired difference in c-index has a 95% CI covering zero, and the automated grade adds no detectable information beyond the clinical grade in a joint model (likelihood-ratio p ≥ 0.27). Adding the non-prostate-specific deaths included in PFI uniformly lowers discrimination. Full tables and figures are in the Supplementary Information.

## 4. DISCUSSION

Applied frozen and uncalibrated to an out-of-distribution, multi-institution cohort and reading a single slide with no pathologist oversight, a gland-level AI diagnostic produced a grade whose standalone recurrence discrimination was not statistically separable from the expert grade. The automated grade’s quadratic Cohen’s kappa falls within the inter-observer range reported for specialist urological pathologists [3] and is broadly consistent with published expert-validated deep-learning graders [8, 9]. Automated grade increased monotonically with clinical grade; the ordering of risk is representative.

Notably, the out-of-distribution generalization described here is measured on the metric that matters for a grading tool rather than on co-acquired internal data that overstates performance through site leakage [12]. Used as the sole predictor, the scenario that actually arises when no pathologist grade is available, the automated grade stratifies recurrence significantly and to within an unresolvable margin of the expert grade (Figures 3, 4). Because that grade is assigned end-to-end from a raw slide with no human annotation, this is direct evidence that grade-based risk stratification can be delivered in no-pathologist settings.

These results do not support a claim of superiority over expert grading. With 24 recurrence events, the study can exclude a large performance gap but not a moderate one, and the comparison is structurally asymmetric: the clinical grade reflects the full prostatectomy specimen while the AI graded a single diagnostic slide, so any residual difference in discrimination conflates genuine grading performance with the tissue available to each system. The clinical grade’s point estimate is higher (c-index 0.78 versus 0.69) and the more sensitive joint test detects a small residual increment (*p* = 0.005). Crucially, this gap is an upper bound on any true grading deficit, because the comparison is structurally asymmetric: the clinical grade reflects the whole prostatectomy specimen, many blocks reviewed by the pathologist, while the AI graded one diagnostic slide (a single slide for 267 of 274 patients). The pathologist therefore had access to tissue, including any higher-grade focus on another block, that the model never saw. A single-slide automated grade matching whole-gland expert discrimination to within a margin this cohort cannot resolve, and subsuming no less of the prognostic signal, is the strong reading of that gap rather than a weak one. The natural test of whether the gap closes, running the model across all available blocks per case, is not possible in TCGA, where one diagnostic slide is the norm, but is feasible in archival cohorts with full block sets.

It is then recommended that the AI prognosis is reported as the continuous high-grade fraction, not the discretized grade group. The continuous readout was the closest proxy to the expert grade and is the more robust target, because the systematic low-end over-call, structural (any trace of pattern 4 promotes 3+3 to 3+4 under area-ranked grading) and amplified by the domain shift between TCGA digitization and the model’s training data [12], distorts the discretized grade more than the continuous one. The over-call concentrates in GG1, precisely where human grading is least reproducible [4] and where the clinical cost of a one-group over-call is lowest. At the high end, the discretized grade under-calls a subset of cribriform-rich high-grade tumors that carry little solid pattern 5; this conflates genuine pattern-5 under-detection with legacy cribriform-grading conventions in the reference and single-slide sampling, and does not extend to tumors where pattern 5 is morphologically florid (which the model grades correctly). Both errors collapse onto grade-group boundaries but largely preserve the underlying high-grade fraction, so a continuous readout, together with cohort-level recalibration of the pattern-4 and pattern-5 decision thresholds, would likely remove most of their impact. We report the uncalibrated behavior deliberately, as the worst case.

The application that motivates the work is harmonization at scale. Large archival and genomically-profiled prostatectomy cohorts, the populations on which genomic classifiers such as Decipher were developed and validated [19, 20], were assembled across many institutions and eras, graded by whichever pathologist reported each case and rarely re-reviewed centrally. An automated grader that matches expert prognostic discrimination out of distribution offers a uniform, reproducible grade across every such slide: consistent grade-based stratification, an automated first-pass over very large slide sets, and a controlled substrate on which to test whether learned embeddings add prognostic value beyond grade, the hypothesis this cohort is too small to settle. In genuinely resource-limited or pathologist-absent settings, the same capability provides a grade-equivalent risk estimate from the slide alone.

## 5. LIMITATIONS

Four patients had unknown pT stage and were imputed to pT2. The cohort has 24 recurrence events; confidence intervals on discrimination are wide, the standalone comparison is powered to exclude a large but not a moderate gap, and the point estimates favor the expert grade. The comparison is also asymmetric in tissue: the clinical grade integrates the whole prostatectomy specimen while the AI graded a single diagnostic slide, so concordance and the residual clinical edge conflate genuine AI grading error with single-slide-versus-whole-gland sampling, biasing the comparison against the model. PSA and surgical margins, the inputs that give clinical nomograms their edge, are unavailable in TCGA, so the grade-plus-stage model is favorable to it; its c-index is an apparent (in-sample) estimate and is correspondingly optimistic. A domain shift between TCGA digitization and the model’s training data produces the documented low-grade over-call; we report uncalibrated performance and applied no stain or scanner normalization. The clinical grade is the original TCGA report, not a central expert re-review, so it is a real-world (variable) reference. The study is single-cohort, retrospective, with ∼32-month median follow-up; the beyond-grade learned-embedding hypothesis is explicitly not tested, as it would overfit at this event count.

## 6. CONCLUSION

From a single out-of-distribution slide, without tuning or a pathologist, a production gland-level prostate AI diagnostic provides standalone disease-free interval stratification not statistically separable from whole-gland expert Gleason grading on TCGA-PRAD and reaches c-index 0.74 when combined with pathologic stage. Our contribution is a validated capability: automated, reproducible, expert-free grade-based risk stratification that generalizes to unseen institutions and radical prostatectomy specimen grading, best delivered as a continuous high-grade fraction, to harmonize and automate grading across the large archival cohorts where uniform expert re-grading is impractical.

## Data Availability

All data produced in the present study are available upon reasonable request to the authors.

## Data and code availability

The cohort definition, the de-identified per-patient feature table, the analysis scripts, and the figure-generation code (which computes every reported c-index and confidence interval) are available in the project repository. Whole-slide images are redistributable from the NCI Genomic Data Commons (TCGA-PRAD); recurrence and clinical endpoints are from cBioPortal (prad_tcga_pan_can_atlas_2018).

## Author contributions

**DDC:** Conceptualization, methodology, formal analysis, data curation, writing — original draft. **JLE:** Software verification, formal analysis, visualization, writing — review and editing. **JS:** Conceptualization, methodology, formal analysis, writing — review and editing. **AP:** Conceptualization, methodology, formal analysis, writing — review and editing.

