## Supporting Information for "An Automated, Pathologist-free Gleason Grade Stratifies Disease-free Interval Comparably to Expert Grading from a Single Out-of-distribution Slide"

Supplementary Information

*Robustness of the prognostic findings across three TCGA-PRAD recurrence endpoints (disease-free interval, progression-free interval, and biochemical [PSA] recurrence).*

### 1. Endpoint definitions

| Endpoint | Event | Source | Note |
| --- | --- | --- | --- |
| DFI (main text) | new-tumor event; deaths censored | prad_tcga_pan_can_atlas_2018 DFS_* (TCGA-CDR) | narrowest disease-recurrence interval |
| PFI | new-tumor event OR death with tumor | prad_tcga_pan_can_atlas_2018 PFS_* (TCGA-CDR) | TCGA-CDR-recommended for PRAD; PFI events ⊇ DFI events |
| BCR (PSA) | post-prostatectomy PSA rise | prad_tcga BIOCHEMICAL_RECURRENCE_INDICATOR + DAYS_TO_* | true prostate-specific signal; legacy study only; few events |

### 2. Prognostic results across the three endpoints

Same frozen PathTools Prostate v11.0 grades and identical analysis (penalizers 0.1 univariate / 0.05 multivariable; B = 2,000 bootstrap; fixed seed); only the survival endpoint changes. The DFI column reproduces the main-text values.

| Quantity | DFI (main text) | PFI | BCR (PSA) |
| --- | --- | --- | --- |
| Patients (n) | 274 | 274 | 234 |
| Recurrence events | 24 | 29 | 12 |
| Median follow-up (months) | 32.0 | 31.5 | 33.1 |
| Quadratic-weighted κ | 0.62 (0.53–0.69) | 0.62 (0.53–0.69) | 0.62 (0.52–0.70) |
| Exact / within-one agreement | 48% / 86% | 48% / 86% | 49% / 86% |
| Univariate Cox — HR (95% CI), p | | | |
| Clinical grade group | 1.44 (1.15–1.79), p=0.001 | 1.41 (1.15–1.74), p=0.001 | 1.30 (1.00–1.68), p=0.050 |
| AI grade group | 1.23 (0.98–1.56), p=0.078 | 1.25 (1.00–1.56), p=0.052 | 1.25 (0.94–1.65), p=0.122 |
| AI %Gleason 4+5 (per SD) | 1.37 (1.03–1.82), p=0.029 | 1.36 (1.04–1.79), p=0.026 | 1.36 (0.97–1.90), p=0.076 |
| AI cribriform area (per SD) | 1.21 (1.01–1.45), p=0.042 | 1.20 (1.00–1.43), p=0.046 | 1.13 (0.85–1.49), p=0.410 |
| Pathologic T stage | 1.90 (1.14–3.15), p=0.014 | 1.64 (1.01–2.66), p=0.047 | 1.70 (0.92–3.14), p=0.093 |
| Standalone discrimination — Harrell c-index (95% CI) | | | |
| Clinical grade group | 0.78 (0.69–0.86) | 0.73 (0.61–0.82) | 0.73 (0.59–0.86) |
| AI grade group | 0.69 (0.58–0.78) | 0.66 (0.55–0.76) | 0.70 (0.55–0.83) |
| AI %Gleason 4+5 | 0.71 (0.60–0.80) | 0.68 (0.57–0.79) | 0.74 (0.61–0.85) |
| AI grade + pT stage (multivariable) | 0.742 (0.63–0.84) | 0.686 (0.56–0.80) | 0.756 (0.58–0.89) |
| Log-rank, 3-group strata — p | | | |
| Clinical grade group | 0.000 | 0.000 | 0.005 |
| AI grade group | 0.022 | 0.029 | 0.031 |
| Equivalence — joint Cox with both grades | | | |
| Joint-model c-index | 0.78 | 0.73 | 0.74 |
| AI adds over clinical (LR p) | 0.500 | 0.429 | 0.273 |
| Clinical adds over AI (LR p) | 0.005 | 0.007 | 0.105 |
| Soft per-tile subset | | | |
| Subset n (events) | 205 (22) | 205 (25) | 193 (12) |
| AI soft mean grade c-index | 0.69 | 0.63 | 0.80 |
| AI soft %Gleason 4+5 c-index | 0.70 | 0.69 | 0.77 |

### 5. Sensitivity figures (PFI and BCR endpoints)

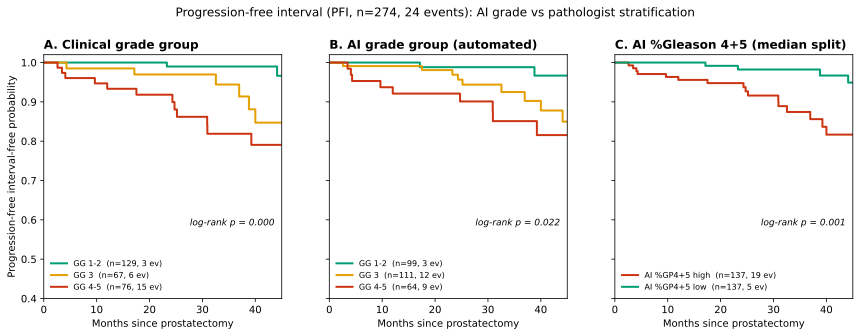

*Figure S1. Kaplan–Meier recurrence stratification on the progression-free interval (PFI) endpoint (n = 274, 29 events). (A) clinical grade group, (B) automated AI grade group, (C) AI %Gleason 4+5 median split; each panel prints its log-rank p.*

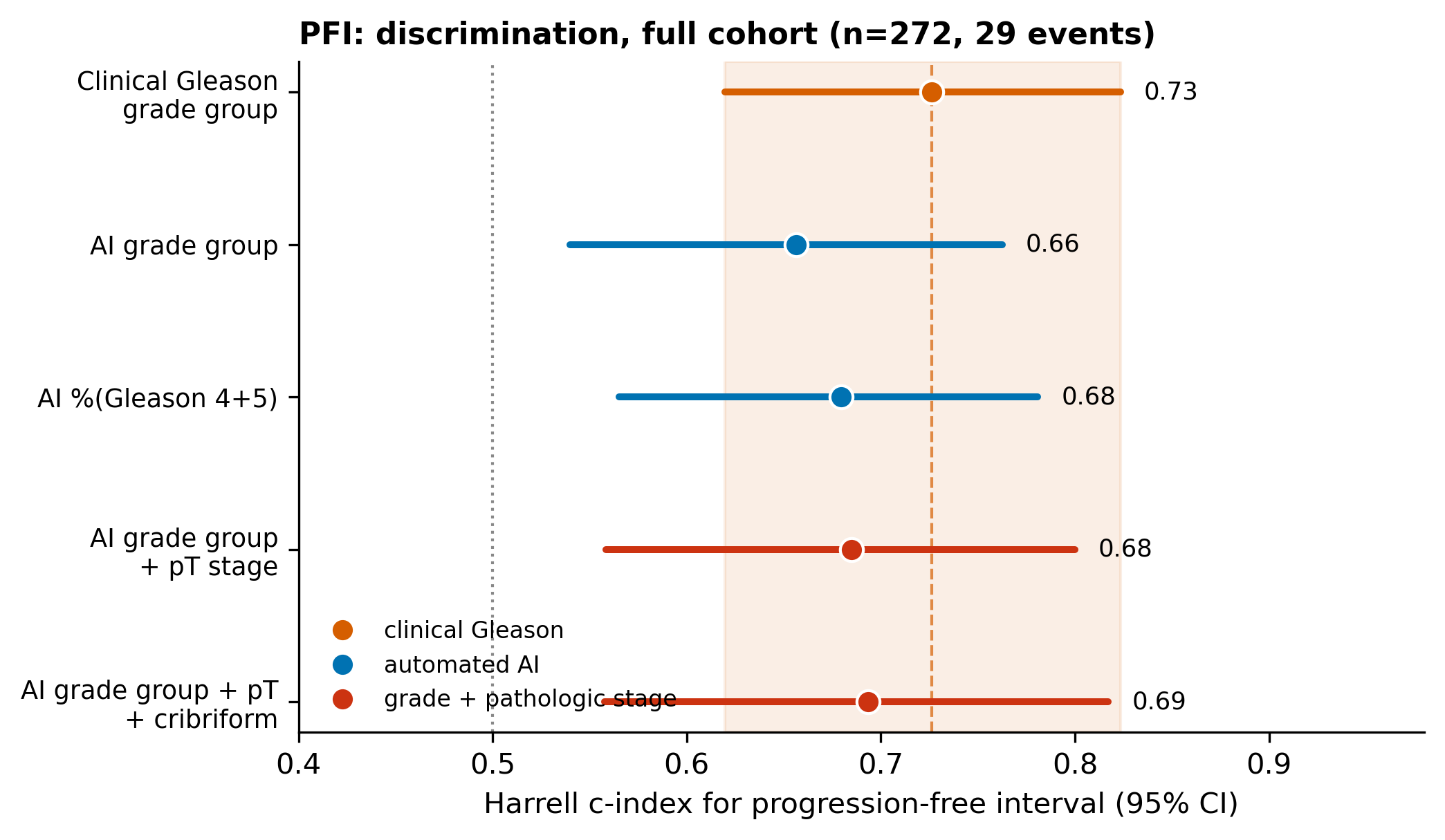

*Figure S2. Standalone c-index (95% bootstrap CI) for clinical versus automated predictors on PFI. The shaded band marks the clinical grade’s CI; every AI predictor overlaps it.*

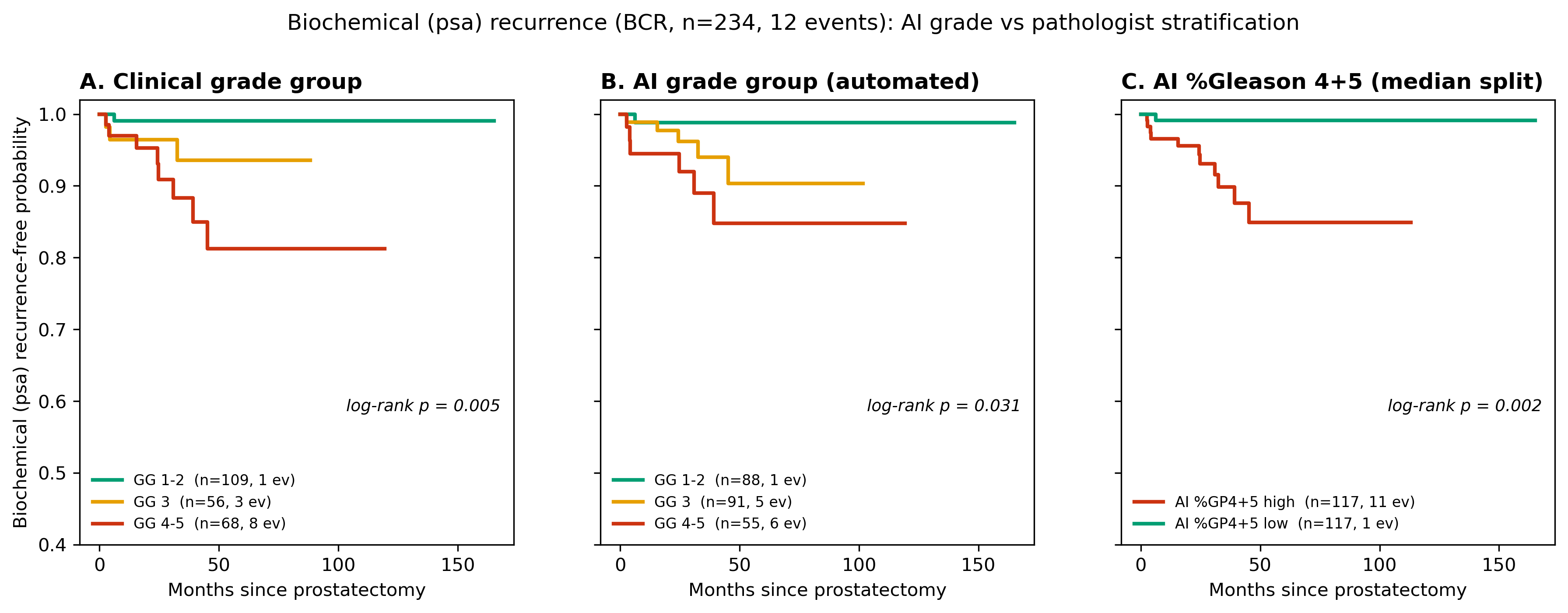

*Figure S3. Kaplan–Meier stratification on true biochemical (PSA) recurrence (BCR; n = 234, 12 events). Panels as in Figure S1. Wide CIs reflect the low event count.*

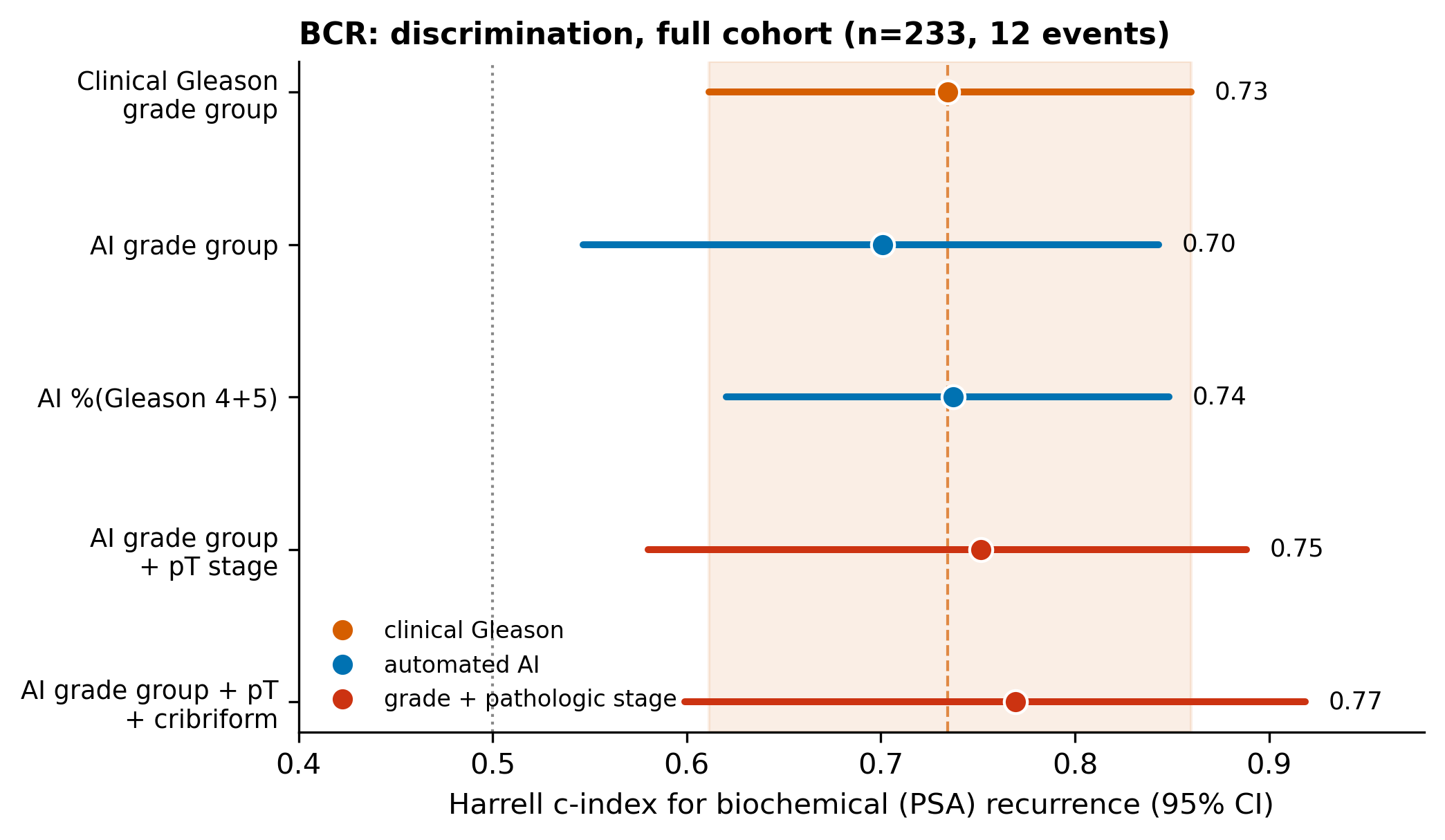

*Figure S4. Standalone c-index (95% bootstrap CI) on BCR. With 12 events the continuous AI %Gleason 4+5 matches the clinical grade and the residual clinical edge is not resolvable; interpret directionally only.*

### 6. Notes

**BCR cohort construction**

Of 274 patients, 234 have a BCR record in the legacy prad_tcga release (16 positive). Twelve BCR-positive patients carry a usable event date (DAYS_TO_BIOCHEMICAL_RECURRENCE_FIRST) and are counted as events; 4 BCR-positive patients lack an event date and are dropped (cannot be placed in time); BCR-negative patients are censored at their DFI follow-up; 36 patients without any BCR record are dropped. The low event count (12) makes BCR estimates imprecise; it is included for completeness as the only prostate-specific endpoint.

**Interpretation**

(1) Concordance is endpoint-independent (κ = 0.62 throughout). (2) Core conclusions are endpoint-robust across DFI, PFI, and BCR. (3) PFI lowers every discrimination estimate by a few points (its added events are non-prostate-specific deaths) and pathologic T stage loses significance. (4) On BCR the residual clinical edge is no longer detectable (clinical-adds LR p = 0.105 vs 0.005 on DFI) and AI %Gleason 4+5 matches the clinical grade, but at only 12 events this is directional. These support retaining DFI as the main-text endpoint with PFI and BCR as sensitivity analyses.
